# Validation of the Teamwork Perceptions Survey in the Operating Theatre

**DOI:** 10.1101/2024.12.03.24318431

**Authors:** Kathryn H. Fahey, Jennifer M. Weller, Jennifer Anne Long

**Affiliations:** University of Auckland, School of Medicine, Centre for Medical and Health Sciences Education

## Abstract

**Background:** Teamwork is recognised as a vital factor in patient safety, and as such we need valid measurement tools to drive change. The aim of this study was to evaluate the validity of the Teamwork Perceptions Survey as a measure of staff perceptions of the functional and relational components of teamwork and leadership in the operating theatre.

**Methods:** The authors developed a 24-item questionnaire, aligned with the goals of a national team training intervention. The Teamwork Perceptions Survey was administered to New Zealand public hospital operating theatre staff before and after a hospital commencing the intervention. These scores were used to explore the psychometric properties of the tool, using Exploratory and Confirmatory Factor Analysis.

**Results:** 2409 (1233 pre- and 176 post-intervention respondents) completed the Teamwork Perceptions Survey. Exploratory factor analysis revealed 3 factors (6 items, communication and shared mental model; 6 items, leadership and role modelling; 5 items, trust and accountability) and resulted in dropping 5 redundant items. The final 17-item, three factor solution was confirmed by confirmatory factor analysis, revealing satisfactory goodness of fit. Cronbach’s alpha was high for the full survey (*α* = 0.947), and each subscale (communication, *α* = 0.887; leadership, 6 items, *α* = 0.911; trust, *α* = 0.872).

**Conclusions:** These data provide evidence to support the validity and reliability of the Teamwork Perceptions Survey as a measure of staff perceptions of teamwork and leadership in the operating theatre. This new measurement tool for the functional and relational components of teamwork and leadership in the operating theatre has the potential for both measuring and driving quality improvement initiatives in teamwork and patient safety.

**Summary:**

- **What is already known on this topic** – Whilst there are a number of established teamwork measurement tools, many completed by an external observer, or do not adequately address the unique operating theatre environment.
- **What this study adds** – This study supports the validity and reliability of a new measure of staff perceptions of the functional and relational components of teamwork and leadership in the operating theatre. It fills a previous gap in available measures in that it is self-completed and is designed for the operating theatre environment.
- **How this study might affect research, practice or policy** – The Teamwork Perceptions Survey has the potential for both measuring and driving quality improvement initiatives in teamwork and patient safety in surgical practice.

---

Teamwork is recognised as a vital factor in patient safety, and as such we need valid measurement tools to drive change.^1^ The validity of a teamwork measurement tool, in general terms, relates to the context and purpose for which it was designed.^2, 3^ A common approach to validity of a measurement tool is content validity-the degree the items are relevant to and representative of the concept and construct validity - how well the items measure the concept.^2^ In this study we consider content validity and the psychometric components of construct validity of a tool designed to measure operating room staff perceptions of teamwork.

The work of operating theatre teams is dynamic, high acuity, and complex, with high interdependency between a group of people with different skills and expertise, all of whom are necessary to do the required work. Excellent teamwork and leadership is necessary in this environment to optimise patient care and avoid unintended harm to patients.^4^ Feeling that you are working well together in a team invokes a sense of belonging, and contributes to the well-being of the team members. This in turn increases the likelihood of the team members engaging in the work of the team, i.e. patient care, and, in the longer term, staying in the health workforce.^5^

## Theoretical Framework

The motivation for this study was to measure changes in operating theatre staff perceptions of teamwork before and after the introduction of the NetworkZ program, a New Zealand-wide multidisciplinary simulation-based team training intervention.^6^ The NetworkZ theoretical model of teamwork draws on the work of Salas and colleagues^7^ who described the underpinning elements observable behaviours demonstrated by high functioning teams. The model prioritises mutual trust and respect, a team orientation versus an individualistic orientation, a desire to create a shared mental model amongst team members, and clear communication strategies. These factors enable teams to be adaptable, because they share information and constantly update team situation awareness^8^ to monitor each other’s behaviours and speak up with concerns, to anticipate the needs of others in the team and offer help, and to embrace inclusive leadership.^9^

We therefore sought a tool designed to measure the perceptions of teamwork by multidisciplinary members of operating theatre teams, and that privileged the relational elements of teamwork and team leadership. Specifically, we required a tool to evaluate operating theatre staff perceptions of teamwork before and after the introduction of a the NetworkZ program.^6^

## Existing Teamwork Measures

To this end, we considered a number of established teamwork measurement tools. Some established tools require external parties or researchers to observe and rate the teamwork skills.^10-13^ However, a measurement tool that is completed by staff rather than researchers extends the value of the tool beyond research, to ongoing quality improvement. Therefore, we sought a self-completed teamwork measurement tool.

A number of self-completed tools have been developed to measure teamwork within a healthcare setting. Some^14-16^ have been developed to measure the *attitudes* of healthcare workers towards teamwork, while our aim was to establish how teamwork was perceived in order to test a team training intervention. The TeamSTEPPS Teamwork Perception Questionnaire (T-TPQ)^17^ is a self-completed teamwork measurement tool that addresses perceptions of teamwork. The T-TPQ was developed to address the four factors that make up TeamSTEPPS team training model and has been validated as a measure of the perceptions of teamwork within healthcare.^18^ However, on review of the T-TPQ items we considered it did not adequately address the unique operating theatre environment, or align with our theoretical framework.^19, 20^

We were unable to identify a tool based on staff perceptions that adequately addressed the components of our theoretical model of teamwork and leadership in the context of the operating theatre. We therefore designed a new tool; the Teamwork Perceptions Survey (TPS). The aim of this study was to explore the validity of the TPS as a measure of staff perceptions of the functional and relational components of teamwork and leadership in the operating theatre. Specifically, we explored item validity through exploratory and confirmatory factor analysis in order to refine our measurement tool.

## Method

### Ethical Approval

Ethical approval was obtained for this study as part of a larger NetworkZ evaluation study. This larger study was approved by the New Zealand Health and Disability Ethics Committee (HDEC; Ref: 16/NTB/143).

### Item Generation Approach

Items were developed using a deductive approach,^2^ through an extensive writing process and by researchers with expertise in teamwork principles and training in a multidisciplinary operating theatre setting. Items were considered in alignment with the goals of the NetworkZ programme which was developed from the theoretical framework proposed by Salas.^7^ The research team generated an initial 40 items, organised into functional, relational, and leadership domains. Consistent with our deductive approach, where applicable we drew on wording from existing, well-validated surveys items.^2^

To remove extraneous items and ensure content validity^2^, the 40-item draft survey underwent iterative review and development by seven clinicians with expertise in team training, and who represented all health professional groups within the operating theatre. This process resulted in 24 items across two domains, relating to senior leadership (8 items) and teamwork behaviours (16 items; Table 1). In addition, the survey collected demographic information on professional group, gender, years of service and ethnicity in order to determine if our respondents were representative of the wider community.

**Table 1.** 24 item Teamwork Perception Survey.

|  |  |
| --- | --- |
| **Senior leader** |  |
| In general, in theatres where I regularly work, senior staff (e.g. consultant surgeon and anaesthetist, senior nurse, charge tech): |  |
| S1 | Provide clear directions to the team. |
| S2 | Use an appropriate balance of assertiveness and support. |
| S3 | Motivate team members to do their best. |
| S4 | Establish a positive atmosphere in theatre care. |
| S5 | Facilitate team problem solving during the case. |
| S6 | Role model acceptable interaction with colleagues. |
| S7 | Engage in pre case briefings of the whole team. |
| S8 | Co-ordinate tasks and individual team member contributions to patient. |
| **Teamwork** |  |
| Team members: |  |
| T9 | Provide support to each other. |
| T10 | Shift work responsibilities to underutilised team members at times of high workload. |
| T11 | Keep an eye on other team members' performance. |
| T12 | Speak up about potential mistakes or lapses in other team members' plans or actions. |
| T13 | Provide constructive feedback regarding team member actions to facilitate self-correction. |
| T14 | Are willing to acknowledge mistakes and accept feedback. |
| T15 | Value the input of all team members to patient care. |
| T16 | Take into account alternative solutions offered by other team members. |
| T17 | Share information about the case planning, progress and concerns with other team members. |
| T18 | Involve the whole team in planning patient care. |
| T19 | Have a good understanding of the roles and abilities of their teammates. |
| T20 | Anticipate and predict each other's needs. |
| T21 | Have a lot of respect for each other. |
| T22 | Enjoy working together. |
| T23 | Exchange information clearly and concisely. |
| T24 | Use names frequently when communicating information or allocating tasks. |

All items were evaluated on a 5-point Likert scale, where 1=“strongly agree” and 5=“strongly disagree”. Items were coded such that a lower score indicated a more positive perception of teamwork.

### Data Collection

This 24-item Teamwork Perceptions Survey (TPS) was administered to New Zealand public hospital operating theatre staff who worked in the five surgical categories included in the NetworkZ national team training program: general, orthopaedic, urology, otorhinolaryngology and plastic surgery. To disseminate the TPS as widely as possible, surveys were distributed in paper and electronic form. The paper survey was handed out directly by a research team member where possible, left in staff mailboxes, and also distributed by local staff in paper form or email. The TPS was distributed in the 3-month period prior to a hospital commencing NetworkZ team training, and again between 15- and 18-months following the training implementation. Staff-lists at each hospital the at time of data collection were used to estimate response rate.

### Data Management

Paper and online surveys were collected by the NetworkZ faculty and entered into an excel spreadsheet by an administrator. Data integrity was ensured by one researcher checking a sample of the data entered in the spreadsheet against original surveys.

### Data Analysis

All responses from Time 1 (pre-intervention time point) were used for the exploratory factor analysis (EFA), and all responses from Time 2 (post-intervention time point) were used for the confirmatory factor analysis (CFA) and reliability testing. This ensured that data for the EFA and CFA were independent.

The Time 1 data set was subject to EFA using maximum likelihood estimation (MLE) with direct oblimin rotation via SPSS Statistics.^21^ The factorial structure obtained using EFA was then subject to CFA on the Time 2 data set, using a structural equation approach via SPSS Amos.^22^ Modification indices (MIs), including additional covariances and unique error terms, were considered to determine the models’ goodness of fit.^23^ To ensure the TPS reliability, responses in the Time 2 data set were assessed for internal consistency via Cronbach’s alpha(α) using SPSS Statistics.^21^

## Results

### Sample Size, Demographics, and Sample Adequacy

A total of 2409 respondents returned completed TPS between December 2016 and June 2022. There were 1233 responses at Time 1 (pre-NetworkZ implementation), and 1176 at Time 2 (post-NetworkZ implementation). Responses came from 25 hospitals across 19 of the 20 District Health Boards (DHBs) which comprised the public health system in New Zealand at the time of the study. Our respondents were multidisciplinary, with a professional role distribution similar to that of the usual staff distribution in the operating theatre. We estimated the response rate to be 37% at Time 1 and 39% at Time 2, based on staff lists of eligible participants at the time of survey completion at each DHB (estimated to be 3760 and 3200 for Time 1 and Time 2 respectively). Demographics were largely consistent across both time points (Table 2).

**Table 2.** Participant Demographics.

|  | **Time 1** |  | **Time 2** |  | **Total** |  |
| --- | --- | --- | --- | --- | --- | --- |
|  | **n** | **%** | **n** | **%** | **n** | **%** |
| **Role** |  |  |  |  |  |  |
| Consultant Surgeon | 165 | 13.4 | 130 | 11.1 | 295 | 12.2 |
| Consultant Anaesthetics | 223 | 18.1 | 176 | 15.0 | 399 | 16.6 |
| Anaesthetic Technician | 176 | 14.3 | 197 | 16.8 | 373 | 15.5 |
| Surgical Registrar/Fellow | 41 | 3.3 | 42 | 3.6 | 83 | 3.4 |
| Anaesthetic Registrar/Fellow | 44 | 3.6 | 38 | 3.2 | 82 | 3.4 |
| Theatre Nurse | 520 | 42.2 | 539 | 45.8 | 1059 | 44.0 |
| Other/Not Reported | 64 | 5.2 | 54 | 4.6 | 118 | 4.9 |
| **Gender** |  |  |  |  |  |  |
| Male | 465 | 37.7 | 439 | 37.3 | 904 | 37.5 |
| Female | 758 | 61.5 | 726 | 61.7 | 1484 | 61.6 |
| Other/Not Reported | 10 | 0.8 | 11 | 0.9 | 21 | 0.9 |
| **Ethnicity** |  |  |  |  |  |  |
| *NZ European | 498 | 40.4 | 578 | 49.1 | 1076 | 44.7 |
| Pacific Peoples | 35 | 2.8 | 31 | 2.6 | 66 | 2.7 |
| Asian Indian | 62 | 5.0 | 88 | 7.5 | 150 | 6.2 |
| Māori | 21 | 1.7 | 35 | 3.0 | 56 | 2.3 |
| Asian | 140 | 11.4 | 226 | 19.2 | 366 | 15.2 |
| Other European | 114 | 9.2 | 139 | 11.8 | 253 | 10.5 |
| Other/Not Reported | 363 | 29.4 | 79 | 6.7 | 442 | 18.3 |
| **Total** | **1233** |  | **1176** |  | **2409** |  |

\*NZ – New Zealand

To ensure the sample was adequate to perform EFA, Keiser-Meyer-Olkin (KMO) and Bartlett’s test of sphericity were considered. We found KMO values of 0.969 for Time 1 and 0.971 for Time 2, suggesting excellent sample adequacy to perform factor analysis. Similarly, Barlett’s test of sphericity returned a significant value for both Time 1, *χ2* (276) = 20089.845, *p* < 0.001, and Time 2, *χ2* (276) = 20194.288, *p* < 0.001. Overall, KMO and Bartlett’s test of sphericity both suggest that the data set is acceptable for factor analysis, and this is consistent across the full data set, Time 1 data set, and Time 2 data set.

### Exploratory Factor Analysis

Using the data set from Time 1 for EFA using maximum likelihood estimation (MLE) with direct oblimin rotation^21^ via SPSS Statistics, different forms of oblique rotation returned consistent solutions.

Contrary to the initially conceptualised two-factor model (i.e., senior leadership and teamwork), Kaiser’s rule (factors with eigenvalues > 1)^24^ suggested a three-factor solution. The suggested three factors cumulatively explained 56.687% of variance (50.243%, 3.546%, and 2.898% respectively).

Initial examinations of factor loadings were somewhat consistent with the anticipated two-factor model (Supplementary Table S1). In line with expectations, items relating to team leadership seemed to load onto factor 2, whilst items relating to teamwork seemed to load onto factor 1. The presence of a third factor was not expected. No unique items loaded on factor 3, however two items relating to a respectful and enjoyable team atmosphere double loaded across factors 1 and 3. Initial factor loadings are shown in Supplementary Table S1.

A number of items showed cross-loading across factors. These were considered in turn, starting with the items with the lowest absolute loading on all factors. If theoretically appropriate, the item was removed and the analysis re-run. The repeat analyses were fixed at three factors, to allow us to consider all possible factors that fit our data at a suitable level (loading > 0.4) with minimal cross-loadings.^24^ Re-running analyses considering eigenvalues over 1 rather than fixed at 3 factors returned consistent solutions. This process was repeated until we established a consistent result with minimal cross-loading.

After considering cross-loadings, a total of four items were removed: S8, T21, T9, and T16 (Supplementary Table S2). Items S8 and T9 showed cross-loading between factors 1 and 2, and items T21 and T16 showed cross-loading between factors 1 and 3. All four items related to multiple factors as opposed to indicating a single underlying factor, both statistically (via cross-loadings) and conceptually. For example, items S8 and T9 could be seen as the responsibility of the leader/senior team-members (factor 1), but also the responsibility of all team members to promote good teamwork (factor 2).

When considering these cross-loadings, we were confident that all four items, whilst relevant to the concept of teamwork we are measuring, were sufficiently covered by other remaining items. For example, item T16 (*take into account alternative solutions offered by other team members*) could be accounted for by item T18 (*involve the whole team in planning patient care*). Therefore, we considered it conceptually valid to remove all four items without compromising the content validity of the TPS. As such, items S8, T21, T9, and T16 were removed.

Following this process, 20 items with no cross-loadings remained, loaded onto three factors. Two items, both loading onto factor 1, returned low factor-loadings below 0.4: T15 (.359), and S7 (.305). We considered these items amongst the other remaining 18, and we felt the model still held theoretical value without these low-loading items. Therefore, beginning with the item with the lowest factor loading (S7), we iteratively removed these items and re-ran the analysis until the solution stabilised.^24^

The resulting analyses revealed a three-factor model with 18 items (Table 3): six items loading on factor 1, seven items on factor 2, and five items on factor 3. This model cumulatively explained 64.962% of variance (52.943%, 7.016%, and 5.003% of variance across factors 1-3 respectively). Items loading on factor 1 related to the instrumental actions to promote teamwork, specifically behaviours designed to promote communication and a shared mental model. Items loading on factor 2 related to the behaviours of the senior team members, relating to leadership and role modelling. Items loading on factor 3 related to the intrinsic trust and accountability required within a team to promote favourable outcomes.

**Table 3.** Final EFA factor loadings.

|  | Item | Factor 1 | Factor 2 | Factor 3 |
| --- | --- | --- | --- | --- |
| T17 | Share information about the case planning, progress and concerns with other team members. | .579 |  |  |
| T18 | Involve the whole team in planning patient care. | .631 |  |  |
| T19 | Have a good understanding of the roles and abilities of their teammates. | .857 |  |  |
| T20 | Anticipate and predict each other's needs. | .735 |  |  |
| T23 | Exchange information clearly and concisely. | .461 |  |  |
| T24 | Use names frequently when communicating information or allocating tasks. | .569 |  |  |
| S1 | Provide clear directions to the team. |  | -.703 |  |
| S2 | Use an appropriate balance of assertiveness and support. |  | -.814 |  |
| S3 | Motivate team members to do their best. |  | -.739 |  |
| S4 | Establish a positive atmosphere in theatre care. |  | -.877 |  |
| S5 | Facilitate team problem solving during the case. |  | -.529 |  |
| S6 | Role model acceptable interaction with colleagues. |  | -.747 |  |
| T22 | Enjoy working together. |  | -.419 |  |
| T10 | Shift work responsibilities to underutilised team members at times of high workload. |  |  | -.509 |
| T11 | Keep an eye on other team members' performance. |  |  | -.581 |
| T12 | Speak up about potential mistakes or lapses in other team members' plans or actions. |  |  | -.665 |
| T13 | Provide constructive feedback regarding team member actions to facilitate self-correction. |  |  | -.925 |
| T14 | Are willing to acknowledge mistakes and accept feedback. |  |  | -.629 |
| Eigenvalues |  | 9.530 | 1.263 | .901 |
| % of variance |  | 52.943 | 7.016 | 5.003 |

**Note.** Only loadings greater than 0.3 displayed.

#### One versus Two versus Three Factor Solutions

Our initial EFA suggested a three-factor model, however when considering the remaining 18-items, only two factors produce eigenvalues above 1, suggesting a two-factor solution may best fit our data. When examining the scree plot, there was a sharp decline following the first point that levels off (Supplementary Figure S1). This suggested a one-factor solution may best-fit our data. Given these multiple possibilities, our CFA initially considered all three potential solutions. However, the three-factor solution consistently showed better model fit compared to the one- and two-factor solutions (Supplementary Table S3). Therefore, we decided to focus on the three-factor solution.

### Confirmatory Factor Analysis

Using the data set from Time 2 for CFA, the initial CFA outcomes suggested adequate goodness of fit, based on a variety of factors (Table 4). Chi^2^ was large and significant, *χ*2 = 1071.797 (132), *p* < 0.000. Given Chi^2^ is dependent on sample size, our large n meant this significance was not unexpected.^24^ The root mean squared error of approximation of the model was significant, RMSEA = 0.78, *p* < 0.000, suggesting the model was statistically significantly close fitting. However, at above 0.7 this could be considered a mediocre fit, with a lower value being preferable and a value below 0.5 ideal.^24^ The goodness of fit indicator, comparative fit index, and Tucker-Lewis index all returned values above 0.9 (GFI = 0.910, CFI = 0.934, TLI = 0.923). For all three measures results above 0.9 is considered acceptable fit, though above 0.95 is considered preferable.^24^

**Table 4.**
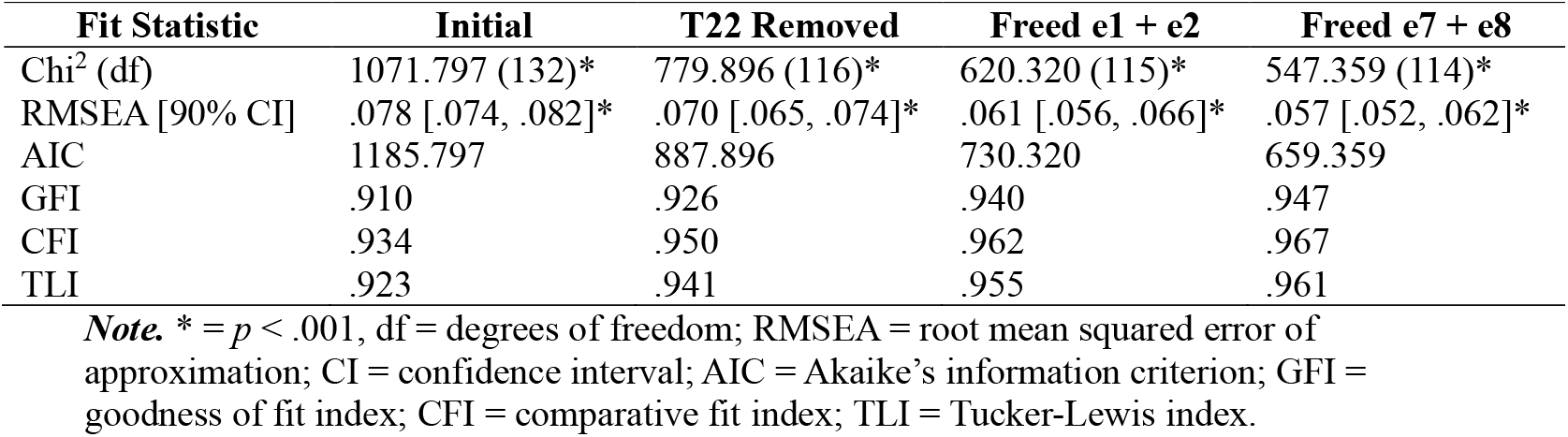
CFA goodness of fit measures across the three-factor model following iterative modification indices.

| Fit Statistic | Initial | T22 Removed | Freed e1 + e2 | Freed e7 + e8 |
| --- | --- | --- | --- | --- |
| $\chi^2$ (df) | 1071.797 (132)* | 779.896 (116)* | 620.320 (115)* | 547.359 (114)* |
| RMSEA [90% CI] | .078 [.074, .082]* | .070 [.065, .074]* | .061 [.056, .066]* | .057 [.052, .062]* |
| AIC | 1185.797 | 887.896 | 730.320 | 659.359 |
| GFI | .910 | .926 | .940 | .947 |
| CFI | .934 | .950 | .962 | .967 |
| TLI | .923 | .941 | .955 | .961 |

**Note.** \* = $p < .001$ , df = degrees of freedom; RMSEA = root mean squared error of approximation; CI = confidence interval; AIC = Akaike’s information criterion; GFI = goodness of fit index; CFI = comparative fit index; TLI = Tucker-Lewis index.

In order to improve goodness of fit, MIs were considered. As MIs are data-driven, there is the possibility that any changes to the model are related only to the data set and thus devoid of theory.^23^ Therefore, when considering MIs, we were careful to examine the proposed modifications within the context of our theoretical framework.

We first considered additional covariances and found that the unique error term (e13) associated with item T22 (“*Enjoy working together*”) had an MI relating to both Factor 1 and Factor 2. We considered that there was some source of variance error relating to this item that may load on both factors 1 and 2, for example, team members may consider enjoying working with their teammates to relate to the communication within the team (items loading on Factor 1), but to also relate to the environment role modelled by the team leader(s) (items loading on Factor 2). We also noted that the EFA suggested item T22 had the lowest factor loading (-.419) of all retained items. Given this theoretical rationale and lower confidence in the factor loading, we eliminated item T22 and re-ran the analyses.

CFA revealed that the goodness of fit for this modified model had improved when compared to the previous model (Table 4), however these were still not within the ideal range. In an effort to further improve goodness of fit measures MIs were again considered, this time investigating the MIs of the unique error terms.

Two correlated-error terms returned very high MIs, above 50. One was for the error values associated with items T17 (“*Share information about the case planning, progress and concerns with other team members*”; error value e1) and T18 (“*Involve the whole team in planning patient care*”; error value e2) loading on Factor 1 (MI = 149.549). The other was for the error values associated with items S1 (“*Provide clear directions to the team*”; error value e7) and S2 (“*Use an appropriate balance of assertiveness and support*”; error value e8) loading on Factor 2 (MI = 67.046).

These reliable correlated-error terms reflect either: 1) unmeasured sources of influence that affect responses to pairs of questions (e.g., situational variation in amount of information or time available to share with the team to aid in planning patient care), or 2) shared sources of error variance (e.g., subjective impressions of what might be considered clear or appropriate from a team leader).

Given the high MIs for these correlated-error terms, and the theoretical link between the items, we decided to re-run the modified model, first freeing the two largest correlated errors (e1 and e2) and then the second (e7 and e8). The final goodness of fit measures suggested good model fit (Table 4). The final factors and associated items are described in Table 5. The confirmed teamwork perceptions 3-factor model is presented in Supplementary Figure S2.

**Table 5.** Confirmed teamwork perception survey factors and items.

| Initial Item Code | Final Item Code | Item and Factor |
| --- | --- | --- |
| **Communication and Shared Mental Model** |  |  |
| T17 | C1 | Share information about the case planning, progress and concerns with other team members. |
| T18 | C2 | Involve the whole team in planning patient care. |
| T19 | C3 | Have a good understanding of the roles and abilities of their teammates. |
| T20 | C4 | Anticipate and predict each other’s needs. |
| T23 | C5 | Exchange information clearly and concisely. |
| T24 | C6 | Use names frequently when communicating information or allocating tasks. |
| **Leadership and Role Modelling** |  |  |
| S1 | L1 | Provide clear directions to the team. |
| S2 | L2 | Use an appropriate balance of assertiveness and support. |
| S3 | L3 | Motivate team members to do their best. |
| S4 | L4 | Establish a positive atmosphere in theatre care. |
| S5 | L5 | Facilitate team problem solving during the case. |
| S6 | L6 | Role model acceptable interaction with colleagues. |
| **Trust and Accountability** |  |  |
| T10 | T1 | Shift work responsibilities to underutilised team members at times of high workload. |
| T11 | T2 | Keep an eye on other team members’ performance. |
| T12 | T3 | Speak up about potential mistakes or lapses in other team members’ plans or actions. |
| T13 | T4 | Provide constructive feedback regarding team member actions to facilitate self-correction. |
| T14 | T5 | Are willing to acknowledge mistakes and accept feedback. |

### Reliability

As a final step, we considered the reliability of our confirmed Teamwork Perceptions Survey. Responses in the Time 2 data set were assessed for internal consistency via Cronbach’s alpha (α) using SPSS Statistics.^21^ Cronbach’s alpha was high for the full survey (17 items, *α* = 0.947), as well as for each individual subscale (communication and shared mental model, 6 items, *α* = 0.887; leadership and role-modelling, 6 items, *α* = 0.911; trust and accountability, 5 items, *α* = 0.872). The internal consistency is generally considered good for Cronbach’s alpha values above 0.8, and excellent for values above 0.9.^25^ Therefore, we conclude that the TPS as a whole, as well as each subscale, is reliable.

## Discussion

Using data from 2409 surveys from multidisciplinary operating theatre staff from covering 95% of the public hospital system in New Zealand, we provide evidence strongly supporting the validity and reliability of the TPS as a measure of staff perceptions of the functional and relational components of teamwork and leadership in the operating theatre. Via EFA we established that our data best fit a three-factor solution, accounting for 64.96% of variance. We confirmed this result via CFA. The final three-factor model included a total of 17 items: six items loading on factor 1 related to the functional components of teamwork, i.e., instrumental actions to promote teamwork; six items loading on factor 2 related to behaviours of the senior team members; and five items loading on factor 3 related to relational components of teamwork, i.e., intrinsic trust and accountability to each other within a team. We labelled the three factors Communication and Shared Mental Model, Leadership and Role Modelling, and Trust and Accountability respectively.

Strengths of our measurement tool include the iterative development and refinement of the items by clinicians from the relevant disciplines with expertise in teamwork, and the strong theoretical framework underpinning our conceptual understanding of the measure, aligning with the work of Salas.^7^ As such we are confident that the TPS encompasses good content validity. Similarly, the verification of the TPS via CFA supports the construct validity of the measure. Furthermore, the high Cronbach’s alpha established for the TPS (both as a complete measure, and for each subscale) supports the reliability of the TPS.^25^ A higher Cronbach’s alpha for the full measure compared to each subscale may be due to each subscale assessing a different aspect of the common concept of teamwork, as well as the higher number of items across the full survey compared to individual subscales.^25^

The TPS adds to the available team measurement tools in a number of ways: it is specifically designed for the context of the operating theatre; it highlights the fundamentally important relational components of teamwork; and it highlights the inclusive elements of leadership, emphasising shared problem-solving. Like a number of other teamwork measurement tools, the items of the TPS provide guidance on desirable attributes of teams at work. In addition, the TPS is a self-report measure and could be used for continuous quality improvement and ultimately contribute to a shift in the culture of the operating theatre. The culture of the operating theatre is often considered steeply hierarchical and lacking in psychological safety and the TPS could contribute to a cultural shift, for example if used by operating theatre departments as part of an intervention to promote reflection by staff on elements for improvement.

### Strength and Limitations

Our study has a number of strengths and limitations. We had large numbers; our respondents were a representative sample of staff from 95% of the DHBs within New Zealand and were representative of the professional groups working in operating theatres. Thus, we can be confident in the generalisability of our measure in operating theatres throughout New Zealand. Due to the common features of operating theatre structure and function in many high-income countries we also suggest this measure may be generalisable to other countries, however this remains to be tested. Our use of MIs to improve model fit is experimental, as the indices are purely data driven.^23^ Therefore, any modifications we made to the model to improve model fit were used sparingly and with valid theoretical reasoning.

Future research could explore the performance of the TPS in different countries or under different conditions in the operating theatre, for example, comparing perceptions of teamwork between ad hoc operating theatre teams (such as in emergency theatres) and staff working in stable operating theatre teams.

## Conclusion

This study provides strong evidence on the validity of the Teamwork Perceptions Survey as a measure of staff perception of the functional and relational components of teamwork and leadership in the operating theatre. The tool comprises 17 items across three factors: communication and shared mental model, leadership and role modelling, and trust and accountability. The TPS is a new measure of the teamwork and leadership in the operating theatre, with the potential for both measuring and driving quality improvement initiatives in teamwork and patient safety.

## Data Availability

All data produced in the present study are available upon reasonable request to the authors

