## Supplemental Figure 1 for "Validation of the Teamwork Perceptions Survey in the Operating Theatre"

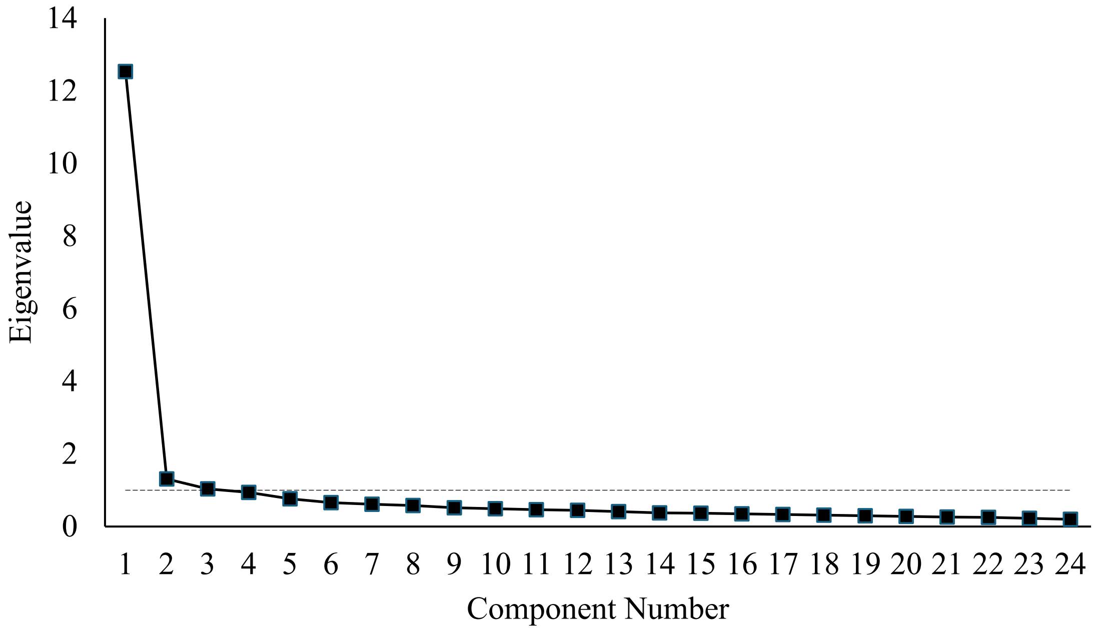


**Figure S1.** *Exploratory factor analysis scree plot.*
***Note.*** Eigenvalues at 1 indicated on figure by dashed line.
