## Supplementary figures and images for "Validation of the Teamwork Perceptions Survey in the Operating Theatre"

### Supplemental Figure 2

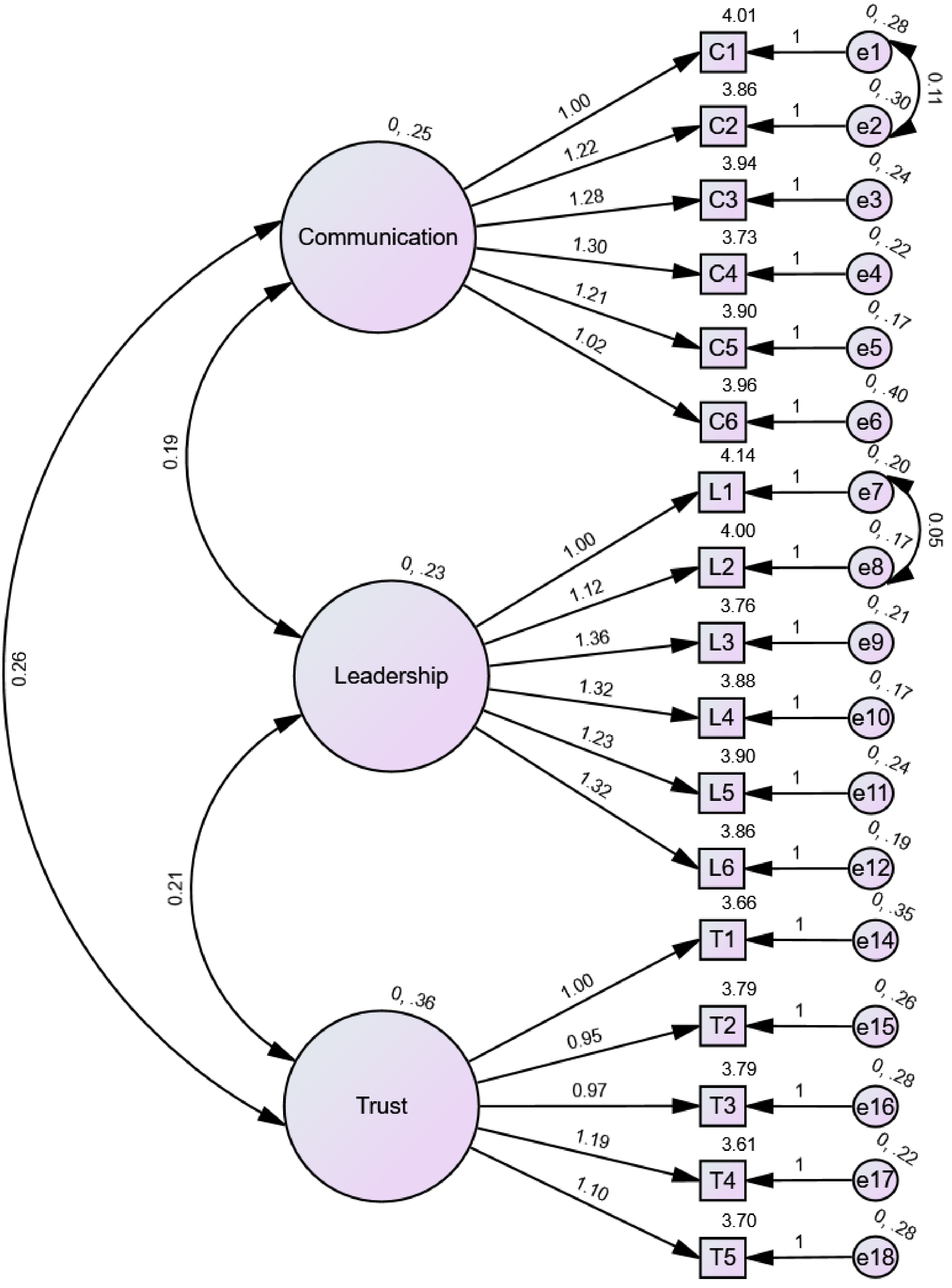


**Figure S1.** *Confirmed teamwork perceptions 3-factor model.*
