## Supplemental Tables for "Validation of the Teamwork Perceptions Survey in the Operating Theatre"

**Table S1**. Exploratory factor analysis factor loadings after rotation.

|  | **Item** | **Factor 1** | **Factor 2** | **Factor 3** |
| --- | --- | --- | --- | --- |
| S1 | Provide clear directions to the team. |  | -0.781 |  |
| S2 | Use an appropriate balance of assertiveness and support. |  | -0.836 |  |
| S3 | Motivate team members to do their best. |  | -0.726 |  |
| S4 | Establish a positive atmosphere in theatre care. |  | -0.804 |  |
| S5 | Facilitate team problem solving during the case. |  | -0.555 |  |
| S6 | Role model acceptable interaction with colleagues. |  | -0.731 |  |
| S7 | Engage in pre case briefings of the whole team. |  | -0.455 |  |
| S8 | Co-ordinate tasks and individual team member contributions to patient. | 0.316 | -0.493 |  |
| T9 | Provide support to each other. | 0.43 |  |  |
| T10 | Shift work responsibilities to underutilised team members at times of high workload. | 0.674 |  |  |
| T11 | Keep an eye on other team members’ performance. | 0.665 |  |  |
| T12 | Speak up about potential mistakes or lapses in other team members’ plans or actions. | 0.747 |  |  |
| T13 | Provide constructive feedback regarding team member actions to facilitate self-correction. | 0.782 |  |  |
| T14 | Are willing to acknowledge mistakes and accept feedback. | 0.744 |  |  |
| T15 | Value the input of all team members to patient care. | 0.592 |  |  |
| T16 | Take into account alternative solutions offered by other team members. | 0.662 |  |  |
| T17 | Share information about the case planning, progress and concerns with other team members. | 0.612 |  |  |
| T18 | Involve the whole team in planning patient care. | 0.595 |  |  |
| T19 | Have a good understanding of the roles and abilities of their teammates. | 0.695 |  |  |
| T20 | Anticipate and predict each other’s needs. | 0.733 |  |  |
| T21 | Have a lot of respect for each other. | 0.356 |  | -0.602 |
| T22 | Enjoy working together. |  |  | -0.587 |
| T23 | Exchange information clearly and concisely. | 0.503 |  |  |
| T24 | Use names frequently when communicating information or allocating tasks. | 0.381 |  |  |
|  | Eigenvalues | 12.527 | 1.307 | 1.038 |
|  | % of variance | 50.243 | 3.546 | 2.898 |

***Note.*** Only loadings greater than 0.3 displayed.

**Table S2.** Order of items removed due to cross-loading.

|  | | | **Cross-Loadings** | | |
| --- | --- | --- | --- | --- | --- |
| **Order Removed** | **Item** | | **Factor 1** | **Factor 2** | **Factor 3** |
| First | S8 | Co-ordinate tasks and individual team member contributions to patient. | .316 | -.493 |  |
| Second | T21 | Have a lot of respect for each other. | .307 |  | -.619 |
| Third | T9 | Provide support to each other. | .338 | -.316 |  |
| Fourth | T16 | Take into account alternative solutions offered by other team members. | .331 |  | -.376 |

**Table S3.** Confirmatory factor analysis goodness of fit statistics across possible models.

| **Fit Statistic** | **1-Factor Model** | **2-Factor Model** | **3-Factor Model** |
| --- | --- | --- | --- |
| *Including Item T22* | | | |
| Chi^2^ (df) | 2258.832 (135)*** | 1414.081 (134)*** | 1071.797 (132)*** |
| RMSEA | .116 (.112, .120)*** | .090 (.086, .094)*** | .078 (.074, .082)*** |
| AIC | 2330.832 | 1524.081 | 1149.797 |
| GFI | .774 | .893 | .910 |
| CFI | .850 | .910 | .934 |
| TLI | .830 | .897 | .923 |
| *Excluding Item T22* | | | |
| Chi^2^ (df) | 2099.500 (119)*** | 1116.987 (118)*** | 779.896 (116)*** |
| RMSEA | .119 (.115, .125)*** | .085 (.080, .089)*** | .070 (.065, .074)*** |
| AIC | 2167.500 | 1220.987 | 853.896 |
| GFI | .773 | .888 | .926 |
| CFI | .849 | .924 | .950 |
| TLI | .828 | .912 | .941 |

***Note.*** * = *p* < .001, df = degrees of freedom; RMSEA = root mean squared error of approximation; AIC = Akaike’s information criterion; GFI = goodness of fit index; CFI = comparative fit index; TLI = Tucker-Lewis index.
